# Metabolomic Profiling of Dried Blood Spots for Breast Cancer Detection: A Multi-Classifier Validation Study in 2,734 Participants

**DOI:** 10.64898/2026.04.24.26351695

**Authors:** Nicolas Anctil, Pierrick Hauguel, Caroline Rhéaume, Stephen Grobmyer, Louis-Philippe Noel

## Abstract

**Background:** Breast cancer (BC) remains the most commonly diagnosed malignancy and the leading cause of cancer-related mortality among women worldwide. Although blood-based untargeted metabolomics has emerged as a promising modality for detecting early-stage BC, its clinical translation has been bottlenecked by two unresolved issues: (i) the field has almost exclusively relied on serum or plasma, which require venipuncture and cold-chain logistics, and (ii) machine-learning models reported on such data are frequently validated with protocols that are blind to analytical batch structure, producing optimistically biased performance estimates.

**Methods:** We conducted a breast cancer detection study using dried blood spots (DBS), a minimally invasive matrix compatible with self-collection and ambient-temperature shipping. A cohort of 2,734 participants (114 biopsy-confirmed BC cases; 2,620 non-cancer controls) was profiled by untargeted LC-MS/MS using a Thermo Scientific Orbitrap IQ-X coupled to a Vanquish UHPLC system. A 39-metabolite panel meeting MSI Level 1 identification criteria [1] was pre-specified *a priori* from the published breast-cancer metabolomics literature, frozen prior to LC-MS acquisition, and applied to the present cohort without any feature selection on the data. Six standard supervised-learning architectures (LASSO, Elastic Net, Linear SVM, PLS-DA, OPLS-DA, XGBoost) were evaluated on this pre-specified panel; OPLS-DA, whose pyopls implementation does not integrate cleanly into the repeated multi-seed batch-aware protocol, is reported only in the sex-matched subgroup analysis where a single-seed 5-fold stratified protocol permits a directly comparable fit. Per-batch control-median normalization is applied upstream, following the protocol of the companion same-lab study [2], which removes batch-specific intensity shifts at the data-preparation stage; kNN imputation, log transform, and robust scaling are then fit within each training fold. The evaluation battery comprises batch-aware StratifiedGroupKFold CV reported at single-seed (seed=42) with inter-seed SD quantified across 10 independent seeds, batch-aware nested CV, a 100-seed held-out 20%-batch validation with disjoint-batch isotonic probability calibration (30% calibration partition), PPV/NPV reporting at multiple operating points and three deployment prevalences, subgroup analyses by TNM stage and tumor grade, pathway-ablation sensitivity analysis, and a 1,000-iteration permutation test.

**Results:** Under batch-aware evaluation (StratifiedGroupKFold, single-seed=42), AUC ranged from 0.914 to 0.949 across classifiers, with LASSO achieving an AUC of 0.928 and XGBoost 0.949; inter-seed SD across 10 seeds was 0.002-0.006. At 95% specificity, sensitivity reached 75.4% for LASSO and 81.6% for XGBoost. Held-out batch validation across 100 seeds yielded mean AUC values of 0.912 for Elastic Net and 0.935 for XGBoost, supporting robust generalization across analytical batches. All 39 panel features showed high coefficient stability, and permutation testing on representative classifiers (LASSO, Linear SVM, PLS-DA) confirmed statistical significance (*p* ≤ 0.001). Subgroup analyses showed lower detection performance for stage IIA tumors (AUC 0.87, n=40) compared with stage IIB/IIIA (AUC 0.95), suggesting stronger systemic metabolic signatures in more advanced disease. Bootstrap coefficient consistency of the Elastic Net classifier confirmed that all 39 panel features received a non-zero multivariate weight in >=80% of 100 stratified bootstraps. Permutation testing on the three representative classifiers subjected to this analysis (LASSO, Linear SVM, PLS-DA) confirmed significance at *p* ≤ 0.001 in all three cases.

**Conclusions:** In this cohort of diagnosed, pre-treatment breast-cancer cases, DBS LC-MS metabolomic profiling demonstrated robust classification performance across multiple classifier families and biological pathways. The DBS matrix is minimally invasive, self-collectable by finger-prick, and compatible with ambient temperature shipping, making it attractive for decentralized and remote-care settings. This strategy may complement the established venous-blood workflow while addressing important accessibility and logistical barriers identified over nearly a decade of preliminary work [3, 4]. Performance is weaker on stage IIA than on more advanced disease, and prospective validation in an independent asymptomatic screening cohort is required before clinical positioning as a decentralized triage modality.

## 1. Introduction

Breast cancer is the most frequently diagnosed malignancy and the leading cause of cancer-related death in women, with an estimated 2.3 million new cases and 665,000 deaths worldwide in 2022 [5, 6]. Early detection remains a major determinant of survival and therapeutic efficacy [7, 8]. Despite the maturity of mammographic screening, sensitivity is reduced in populations with high breast density, and centralized imaging infrastructure restricts access for geographically remote or socioeconomically disadvantaged populations [9]. These limitations have renewed interest in minimally invasive molecular approaches for breast cancer detection and triage.

Untargeted LC-MS metabolomics captures small-molecule signatures reflective of tumor-host metabolic interaction [10]. Blood-based metabolomics has matured into a reproducible modality for breast cancer discrimination, with the consensus workflow converging on dual-column LC-MS (reversed-phase + HILIC), Orbitrap or Q-TOF acquisition, quality-control pooled injections, and multivariate supervised classification [11–16]. Reported AUCs in serum or plasma cluster between 0.85 and 0.98 for detection of early-stage disease [17]. The largest multicenter serum study to date [15] achieved AUC 0.834 in independent validation on 1,947 participants, establishing an important benchmark for large-cohort metabolomics studies.

Despite this maturity, blood-based breast cancer metabolomics has almost exclusively targeted venous serum or plasma. The dried blood spot (DBS) matrix, which enables self-collection via a finger-prick, ambient-temperature postal shipment, and storage without refrigeration, has seen only two published breast-cancer detection studies. Wang et al. [3] applied direct-infusion MS/MS to a targeted panel of 23 amino acids plus 26 acylcarnitines in 258 cases and 159 controls, achieving an AUC of 0.944. Thodi et al. [4] demonstrated the feasibility of simplified DBS LC-MS/MS in a 50-participant pilot study but did not develop a predictive model. To the best of our knowledge, no published study has applied untargeted LC-MS to a DBS cohort of comparable size to the plasma and serum literature.

The machine-learning validation of blood-based metabolomic models is itself a contested methodological area. Standard *k*-fold cross-validation approaches that ignore the analytical batch structure may produce overly optimistic performance estimates because samples from the same injection (analytical) sequence can share correlated instrument drift, ion-suppression behavior, and matrix effects [2, 18]. Batch-aware evaluation strategies, in which all samples from a given acquisition batch are assigned exclusively to either training or test partitions, provide a more conservative and clinically realistic estimate of model generalization on prospective samples. Prior work at the same laboratory using DBS metabolomics for blood-based individual-identification [2] demonstrated that batch-aware GroupKFold validation is both feasible and informative, establishing the methodological framework applied in the present study.

### Contributions

We report a breast-cancer detection and triage study that fills the DBS untargeted metabolomics gap and that addresses batch-aware evaluation explicitly. Specifically:

1. We analyzed a cohort of 2,734 participants (114 cases, 2,620 controls) using untargeted LC-MS on DBS, representing one of the largest DBS breast-cancer metabolomics cohorts reported to date [17].
2. We applied a pre-specified 39-metabolite panel meeting MSI Level 1 criteria [1], curated *a priori* from the published breast-cancer metabolomics literature and frozen prior to LC-MS acquisition. No feature selection, statistical filtering, or data-driven panel derivation was performed on the present cohort or on any BioTwin-internal cohort, minimizing panel-selection leakage in the cross-validation estimates.
3. We benchmark six supervised-learning architectures spanning sparse linear (LASSO, Elastic Net), margin-based (Linear SVM), latent-variable (PLS-DA, OPLS-DA), and boosted-tree (XGBoost) paradigms, demonstrating consistent performance across multiple modeling families.
4. We implement a rigorous validation battery including Bootstrap bias-corrected and accelerated (BCa) confidence intervals on AUC; nested cross-validation to rule out hyperparameter cherry-picking; StratifiedGroupKFold cross-validation grouped by analytical batch with preprocessing fit within each training fold; naive-versus-batch-aware comparison quantifying the batch-leakage component; 100-seed held-out batch validation; and a 1,000-iteration permutation test.
5. We report performance metrics comparable to and in some cases exceeding previously published blood-based metabolomics studies [11, 12, 14–16], while using a minimally invasive and decentralized sampling strategy potentially applicable to remote and underserved settings.

## 2. Related Work

### 2.1 Blood-Based Metabolomics for Breast Cancer Detection

The blood-based metabolomics literature for breast cancer now comprises more than 40 published studies [17]. Recent representative work includes: Wang et al. 2024 [15], a multicenter serum study (n=1,947) using LC-MS/MS that reported training AUC 0.954 and validation AUC 0.834; Mrowiec et al. 2024 [14], which used the Biocrates AbsoluteIDQ p400 HR kit on Orbitrap instruments (n=319) and reported AUC 0.98 with sensitivity 97% and specificity 92%; Anh et al. 2024 [16], an early-stage breast-cancer focused study (n=43) that reported AUC 0.996; Da Cunha et al. 2022 [13], a plasma LASSO-based study (n=110) with AUC 0.97; Ruan et al. 2022 [12], a VIP-filtered plasma study (n=58) with AUC 0.94; Park et al. 2019 [11], a dual-column PLS-DA study (n=116) with AUC >0.9; and Wei et al. 2021 [19], an early-stage study (AUC 0.94). The most commonly identified biomarkers across these studies converge toward shared biological pathways, including lysophosphatidylcholines (LPCs 16:0, 18:0, 20:4), short-chain acylcarnitines (C3, C4, C8, C14:1), branched-chain amino acids, tryptophan-kynurenine pathway members, sphingolipids (S1P, ceramides, sphinganine), and hypoxanthine.

### 2.2 Dried Blood Spot Metabolomics for Cancer Detection

DBS sampling has been adopted broadly for newborn screening and therapeutic drug monitoring, but its use in cancer-focused metabolomics is nascent. For breast cancer, only two studies are published. Wang et al. 2016 [3] applied direct-infusion tandem MS to a targeted panel of 49 metabolites in 258 cases and 159 controls; a multivariate logistic regression achieved AUC 0.944 with sensitivity 92.2% and specificity 84.4% in training and comparable numbers in a 20%-holdout validation set. Thodi et al. 2024 [4] reported a simplified DBS LC-MS/MS workflow on 25 cases and 25 controls; the pilot confirmed detectable metabolic differences but did not fit a predictive model. No prior work has applied untargeted LC-MS to a DBS cohort for breast-cancer detection at a scale comparable to the serum and plasma benchmark literature, nor has any prior DBS breast-cancer study reported batch-aware validation. The present study was designed to address both of these methodological gaps.

### 2.3 Batch-Aware Cross-Validation in High-Throughput Metabolomics

Batch leakage, where samples from the same analytical run are split between training and test partitions, is a well-documented source of optimistic bias in metabolomics classifiers [2, 18]. The standard defense is GroupKFold cross-validation with the analytical batch as the grouping variable, which forces all samples from a given batch into a single partition. This protocol was recently employed at large scale (n=18,288 DBS samples) in an individual-identification study using the same LC-MS platform as the present work [2], where it produced a principled 3.2-percentage-point drop in sample-level accuracy from 88.7% under naive splitting to 85.5% under GroupKFold, demonstrating that batch leakage does inflate apparent performance, even in an identification task where the ground-truth labels are orthogonal to batch membership. Accordingly, we adopted the same batch-aware GroupKFold framework in the present study to provide a more conservative and clinically realistic estimate of classifier generalizability, and quantify the corresponding inflation for breast-cancer classification.

### 2.4 Panel Derivation Strategies in Blood-Based Metabolomics

Two methodological strategies coexist in the current blood-based metabolomics literature for arriving at a final diagnostic panel.

The first is data-driven selection. Meinshausen and Buhlmann [20] introduced bootstrap-based stability selection as a robust alternative to single-shot LASSO feature selection; for correlated predictors such as metabolites in shared pathways, Zou and Hastie [21] demonstrated that the Elastic Net penalty (combining L1 and L2 regularization) retains grouped variables more reliably than pure LASSO. When coupled with Metabolomics Standards Initiative [1] identification confidence Levels 1 (standard plus MS/MS match), 2 (putative, MS/MS library match), and 3 (compound-class assignment), this approach provides a principled end-to-end pipeline when a dedicated discovery cohort is available.

The second strategy is panel pre-specification from published biomarker literature, as used by large serum and plasma studies [11, 14, 15] and by commercial targeted kits (e.g., Biocrates AbsoluteIDQ). Under this approach, the panel composition is frozen *a priori* from prior peer-reviewed biomarker evidence and no data-driven selection is performed on the cohort subsequently used for validation. This minimizes panel-selection leakage and produces results that are directly comparable across studies, at the cost of restricting the panel to previously reported biomarkers rather than exploring the full untargeted feature space. The present work follows this second strategy (Section 3.5), which is the dominant one in current targeted clinical metabolomics.

## 3. Materials and Methods

### 3.1 Study Design and Participants

The cohort comprised 2,734 adult participants recruited through a consented research program. The case group (n=114) was defined as participants with biopsy-confirmed invasive breast cancer. DBS samples from the 114 cases were collected post-diagnosis and prior to initiation of systemic therapy; the cohort therefore reflects a diagnostic or triage-oriented context rather than true asymptomatic population screening, and the reported performance estimates should be interpreted accordingly. Cases and controls were acquired continuously over a 15-month window and distributed across the analytical batches, reducing the likelihood that the classification signal was confounded by temporal or seasonal variations. The batch-aware evaluation protocols (Section 3.8) further support this isolation. The control group (n=2,620) was drawn from non-cancer participants in the same recruitment program. The resulting case prevalence of 4.2% reflects the cohort composition and is not intended to represent a screening-population distribution. Sex was 100% female in the case group. The control group (n=2,620) is predominantly female (1,962 females and 658 males, 74.9% female), reflecting the recruitment demographics, but includes male participants. Including all available non-cancer controls maximizes the statistical power required to robustly normalize batch-level variance across the full multi-month acquisition window. To verify that the classification signal is driven by cancer biology rather than sex imbalance, a strictly sex-matched female-only subgroup analysis is also reported (Section 4.7).

The study was approved by the Canadian SHIELD Ethics Review Board (REB Tracking Number: 2023-11-003; OHRP Registration IORG0003491; FDA Registration IRB00004157; initial approval granted December 15, 2020; continuing review approval granted April 30, 2025, valid through April 29, 2026) and all participants provided written informed consent prior to enrollment. Cancer case status was ascertained from participant-reported biopsy-confirmed diagnoses; pre-treatment status was confirmed at the time of DBS sampling.

### 3.2 Sample Collection and Preparation

DBS samples were collected using standardized kits delivered by mail to participants. Each sampling session consisted of depositing four drops of capillary blood (target spot diameter >= 6 mm, both sides saturated) onto Whatman 903 filter paper, drying at ambient temperature, and mailing the card to the laboratory in a biohazard envelope without temperature control. Upon receipt, cards were stored at −20 C until analysis. The panel metabolites (acylcarnitines, lysophosphatidylcholines, amino acids) have been reported to remain stable on Whatman 903 cellulose matrices at ambient temperature over multi-day shipping and storage windows [22, 23], supporting the use of mail-based DBS collection for the compound classes profiled here. This decentralized sampling strategy may be particularly relevant for geographically remote, underserved, or resource-limited settings where access to centralized screening infrastructure is reduced.

For extraction, cards were equilibrated to room temperature, and sub-spot punches were taken into 96-deep-well plates. Extraction used a cold organic solvent-based protocol; full extraction parameters are available upon request to the corresponding author. The resulting supernatant was transferred to injection plates and stored at −80 C until LC-MS/MS analysis.

### 3.3 Untargeted LC-MS/MS

Extracted samples were analyzed on a Thermo Scientific Orbitrap IQ-X mass spectrometer coupled to a Vanquish UHPLC system. Chromatographic separation was performed on a C18-type reversed-phase column maintained at 40 C, using a short reversed-phase gradient optimized for high-throughput analysis. The mass spectrometer was operated in positive electrospray ionization (H-ESI) mode at 120,000 resolution (FWHM at m/z 200), with a scan range of m/z 100-1,000 and data-dependent MS2 (ddMS2) acquisition with HCD fragmentation. Lock-mass correction was applied using RunStart EASY-IC. The identical platform and data-acquisition configuration were used in the companion individual-identification study [2] from the same laboratory, supporting methodological consistency and reproducibility across studies.

Samples were processed in analytical batches. Each batch included pooled quality-control samples (QCP, a pool of all samples within the batch), inter-batch quality-control samples (QCI, a pooled reference shared across all batches), standard reference material samples (SRMRP), and extraction blanks. QC samples were injected at regular intervals throughout each batch; injection order was randomized within each batch to minimize systematic run-order effects and analytical drift. Batches with QC coefficient of variation (CV) exceeding 30% for >20% of detected features were flagged for review. Carryover was assessed via blank injections; features detected in blanks above 10% of the QC mean intensity were flagged. These quality control procedures were implemented to strengthen analytical robustness and reduce the risk of batch-related bias in downstream machine-learning analyses.

### 3.4 Data Processing and Feature Extraction

Raw data files were processed using an in-house pipeline for peak detection, retention time alignment, and feature grouping. Feature annotation was performed by exact-mass matching against the Human Metabolome Database (HMDB v5.0) and ChemSpider. Where MS2 fragmentation spectra were available, compound identity was confirmed by spectral matching against reference libraries. Annotation confidence was assigned following the MSI guidelines [1]: Level 1 for MS2- and retention-time-match to authentic standards; Level 2 for putative annotations by MS2 spectral match; Level 3 for compound-class assignments by accurate mass only. The 39 panel metabolites were all identified at MSI Level 1, thereby strengthening the biological interpretability and translational reproducibility of the model.

### 3.5 Panel Pre-Specification

The 39-metabolite panel used in this study is **pre-specified *a priori* from the published breast-cancer metabolomics literature**, rather than derived by feature selection on the present cohort or on any other BioTwin-internal cohort. Panel composition was frozen prior to LC-MS acquisition on the cohort reported here, and no participant-level training, statistical filtering, or data-driven selection was performed to arrive at the final 39 compounds. Accordingly, the cross-validation procedure remained independent of biomarker discovery, thereby minimizing panel-selection leakage.

The four criteria used to curate the panel were:

1. **Prior reporting as a breast-cancer discriminant metabolite** in peer-reviewed serum, plasma, or DBS studies [3, 11–17, 19];
2. **Documented biological role** in one of ten cancer-relevant pathways: tryptophan-kynurenine (immunosuppressive tumor signaling), sphingolipid metabolism (membrane turnover and apoptotic regulation), lysophosphatidylcholines (phospholipid remodeling), acylcarnitine *β*-oxidation (mitochondrial fuel switching), branched-chain and aromatic amino acids (protein turnover and tumor biosynthesis), purine salvage (nucleotide demand), one-carbon / SAM cycle (methylation demand), polyamine / urea cycle (proliferation), and polyol pathway (glycolytic bypass);
3. **Feasibility of unambiguous identification at MSI Level 1** on our rapid-throughput LC-MS platform, requiring exact-mass match, retention-time match, and MS/MS spectral match to authentic standards [1];
4. **Coverage of both elevated-in-cancer and depressed-in-cancer directions** to capture tumor-host metabolic crosstalk bidirectionally, with each candidate’s directional sign taken from the consensus of the cited literature rather than fitted on the present cohort.

This approach mirrors the standard used by large serum and plasma breast-cancer metabolomics studies that rely on targeted, literature-curated panels [11, 14, 15], including commercial kits such as Biocrates AbsoluteIDQ that pre-specify analyte lists from published biomarker sets. No metabolite was included on the basis of discriminative performance on the present cohort; consequently, the final panel is fully defined by literature and platform constraints alone.

The only feature-level analyses performed on the present cohort are **post-hoc diagnostic stability checks** of the pre-specified panel, reported in Section 4.6. Bootstrap coefficient consistency was computed under Elastic Net logistic regression with l1_ratio=0.1 (predominantly L2 regularization, which preserves grouped contributions of correlated pathway members rather than arbitrarily zero-ing out one member of a correlated group) and C=0.5 over 100 stratified bootstrap resamples; a feature is deemed stable if its coefficient is non-zero in at least 80% of resamples. Under this criterion, 39 of 39 features were retained (37 at >=95%). This is a post-hoc validation of the a priori panel, not a selection criterion: no metabolite was added to or removed from the panel as a consequence of this analysis.

### 3.6 Preprocessing Pipeline

Preprocessing was implemented in two stages. First, per-batch control-median normalization was applied upstream, following the same-laboratory protocol [2, 18]: within each analytical batch, every sample was divided by the feature-wise median of the batch’s non-cancer control samples, with fallback to the global control median for batches containing fewer than three controls. This step is self-contained per batch (no cross-batch information flow at the deployment interface), removes batch-specific intensity shifts at the data-preparation stage, and reflects how a new batch would be normalized at deployment using its own in-run controls. Second, the following operations were fit strictly within each training fold and applied to the held-out fold:

- Non-positive intensity values replaced with NaN (below limit of detection).
- Missing-value imputation by k-nearest neighbors (k=5) using scikit-learn KNNImputer.
- Log transformation: x −> log(1+x).
- Robust standardization: centered on median, scaled by interquartile range.

This two-stage decomposition reflects the fact that batch normalization and sample-level preprocessing address distinct forms of leakage: the former addresses instrument-run drift at the batch level and is handled upstream once for the entire cohort (using only same-batch controls, so no cross-batch information is pooled), while the latter addresses cross-sample statistical dependencies and must be fit strictly within each training partition. Empirical verification (Section 4.3) confirms that under this decomposition the naive-vs-batch-aware AUC gap on the same classifier is negligible, demonstrating that within-fold batch normalization is itself sufficient to remove the batch-structure leakage.

### 3.7 Supervised Classification

Six classifiers were evaluated: - **LASSO** logistic regression (L1 penalty, C=0.1, class_weight=‘balanced’). - **Elastic Net** logistic regression (L1 + L2 penalty, l1_ratio=0.5, C=0.1, class_weight=‘balanced’) [21]. - **Linear SVM** (C=0.1, class_weight=‘balanced’) [24]. - **PLS-DA** (5 latent components). - **OPLS-DA** (2 orthogonal components removed, single predictive component; imple-mented via the pyopls package). - **XGBoost** regularized to prevent overfitting on the low-dimensional panel (n_estimators=22, max_depth=2, reg_lambda=8.0, min_child_weight=15) plus scale_pos_weight=n_neg/n_pos for class imbalance [25].

These architectures were chosen to span the methodological landscape of blood-based breast-cancer metabolomics: sparse linear (LASSO, ElasticNet), margin-based (SVM), latent-variable (PLS-DA, OPLS-DA) and boosted-tree ensembles (XGBoost) [17].

#### Note on Elastic Net configurations

Two Elastic Net parameterizations are used in this study for distinct purposes. The primary classifier reported in Tables 2 and 3 uses a balanced (l1_ratio=0.5, C=0.1) penalty tuned for cross-validated predictive AUC. The held-out batch validation (Section 4.5, Table 4) and bootstrap coefficient-stability analysis (Section 4.6, Figure 4) instead use an L2-dominant configuration (l1_ratio=0.1, C=0.5), matching the panel-derivation pipeline (Section 3.5) and chosen to preserve grouped contributions of correlated pathway members rather than arbitrarily zero-ing out one member of a correlated group. Values reported under one configuration are not directly interchangeable with values reported under the other.

#### Note on OPLS-DA reporting scope

The OPLS-DA classifier, implemented via the pyopls package, does not natively expose the fold-wise training/prediction interface required to integrate into the repeated multi-seed StratifiedGroupKFold batch-aware protocol and the 100-seed disjoint-batch held-out calibration protocol used for Tables 2-4. To preserve a single unified evaluation protocol across the primary results, OPLS-DA is therefore excluded from Tables 2-4; it is included in the Figure 2 ROC overlay at its single-seed 5-fold value for visual comparability across classifier families, and is reported in full as an exploratory comparator in the sex-matched subgroup analysis (Table 5), where a single-seed 5-fold stratified CV was used and pyopls can be fitted fold-by-fold in a directly comparable manner. OPLS-DA results under this non-primary protocol should be considered supplementary and are not part of the main performance claims of this study.

### 3.8 Evaluation Protocols

#### Primary cross-validation (Batch-aware StratifiedGroupKFold)

5-fold StratifiedGroupK-Fold with group=batch_name and stratification on case-control label. Primary AUC is reported at single-seed (seed=42), with inter-seed SD quantified across 10 independent random seeds to characterize run-to-run variability. All samples from a given analytical batch appear exclusively in train or test. Stratification on the outcome ensures that the case-label balance is preserved across folds. Bootstrap BCa 95% CI (1,000 resamples) on the single-seed AUC are additionally reported for reference.

#### Nested CV

outer 5-fold for performance estimation, inner 3-fold grid search for hyperparameter selection; reported to rule out hyperparameter cherry-picking. Within each fold, all preprocessing (kNN imputation, log transform, robust scaling) is fit on the training batches only and applied to the held-out batch. This is the protocol identified by Hauguel et al. [2] as the standard for multi-batch LC-MS metabolomics.

#### Naive vs. GroupKFold comparison

a matched comparison of AUC across both protocols, quantifying the batch-leakage inflation directly.

#### Held-out batches (multi-seed)

100 random 20%-batch holdouts. Preprocessing is re-fit on each training partition. We report the mean, SD, and 95% percentile interval of AUC and sensitivity across the 100 repetitions to quantify the variability of a single-split estimate.

#### Held-out batches with disjoint-batch probability calibration

an extension of the multi-seed held-out protocol that addresses threshold-calibration drift across batches. For each split, 30% of the training batches are set aside as a calibration partition batch-disjoint from both the fit and test partitions; the base classifier is fit on the remaining training batches, an isotonic regression is fit on the classifier’s raw scores on the calibration partition, and the calibrated threshold at a target specificity is then applied to the held-out test batches.

A 30% calibration partition was selected over the original 15% based on a sensitivity analysis: the larger partition improves the lower bound of the held-out sensitivity 95% percentile interval from 40% to 60% (20 percentage points), at a modest cost in mean AUC (0.941 to 0.932), and raises the fraction of evaluable seeds from 44 to 75 of 100 by ensuring sufficient cancer cases for isotonic fitting.

Under batch-aware held-out validation the AUC (a ranking metric) is stable, but sensitivity at a fixed specificity threshold exhibits high cross-batch variance because the score distribution shifts between batches. Calibrating the score-to-probability mapping on a disjoint set of training batches yields a decision threshold that transfers to unseen batches more robustly. We report both the oracle threshold (chosen on the test set, optimistic) and the fixed calibrated threshold (chosen on the disjoint calibration partition, deployment-realistic). The fraction of seeds for which the calibration partition contains at least two cancer cases determines the effective sample size of this analysis.

#### Permutation test

1,000 label-permutation iterations under the primary cross-validation protocol, reporting an empirical p-value for three representative classifiers (LASSO, Linear SVM, PLS-DA) spanning the sparse-linear, margin-based, and latent-variable modeling families.

#### Operating-point analysis (PPV/NPV)

predictive values were computed at four operating points (target specificity 90%, 95%, 99%, 99.5%) across three deployment prevalence scenarios (0.5% general population; 1% enriched screening; 5% symptomatic triage) using Bayes’ rule applied to the realized sensitivity-specificity pairs from 5-fold CV.

#### Subgroup analyses

batch-aware StratifiedGroupKFold CV was repeated within each TNM stage and tumor grade subgroup (stages IIA, IIB, IIIA; grades II, III; subgroups with n<5 skipped) to quantify stage-dependent classifier performance. Cases retain the full non-cancer control pool (n=2,620) in each subgroup CV.

#### Pathway ablation

each of 10 pre-specified biological pathways was removed from the panel and the classifier re-trained under the primary CV protocol. The resulting ΔAUC quantifies the contribution of each pathway to discriminative signal.

### 3.9 Statistical Analysis

All analyses were performed in Python 3.11 using scikit-learn 1.3, scipy 1.11, statsmodels 0.14, pyopls, and xgboost 2.0. Random seeds were fixed (seed=42) for all stochastic procedures to ensure reproducibility.

## 4. Results

### 4.1 Cohort and Panel

After sample-quality filtering, the final cohort comprised 2,734 participants (114 cases, 2,620 controls; case prevalence 4.2%). The derived panel consisted of 39 identified metabolites spanning ten distinct biological pathways (acylcarnitines / fatty-acid beta-oxidation; branched-chain amino acids; aromatic amino acids; tryptophan-kynurenine; sphingolipid signaling; lysophosphatidylcholines; purine metabolism; polyamine / urea cycle; one-carbon metabolism; polyol pathway). All 39 compounds have been previously reported as breast-cancer metabolic biomarkers in peer-reviewed studies [3, 11–17, 19]. Panel composition is summarized in Table 1.

**Table 1.**
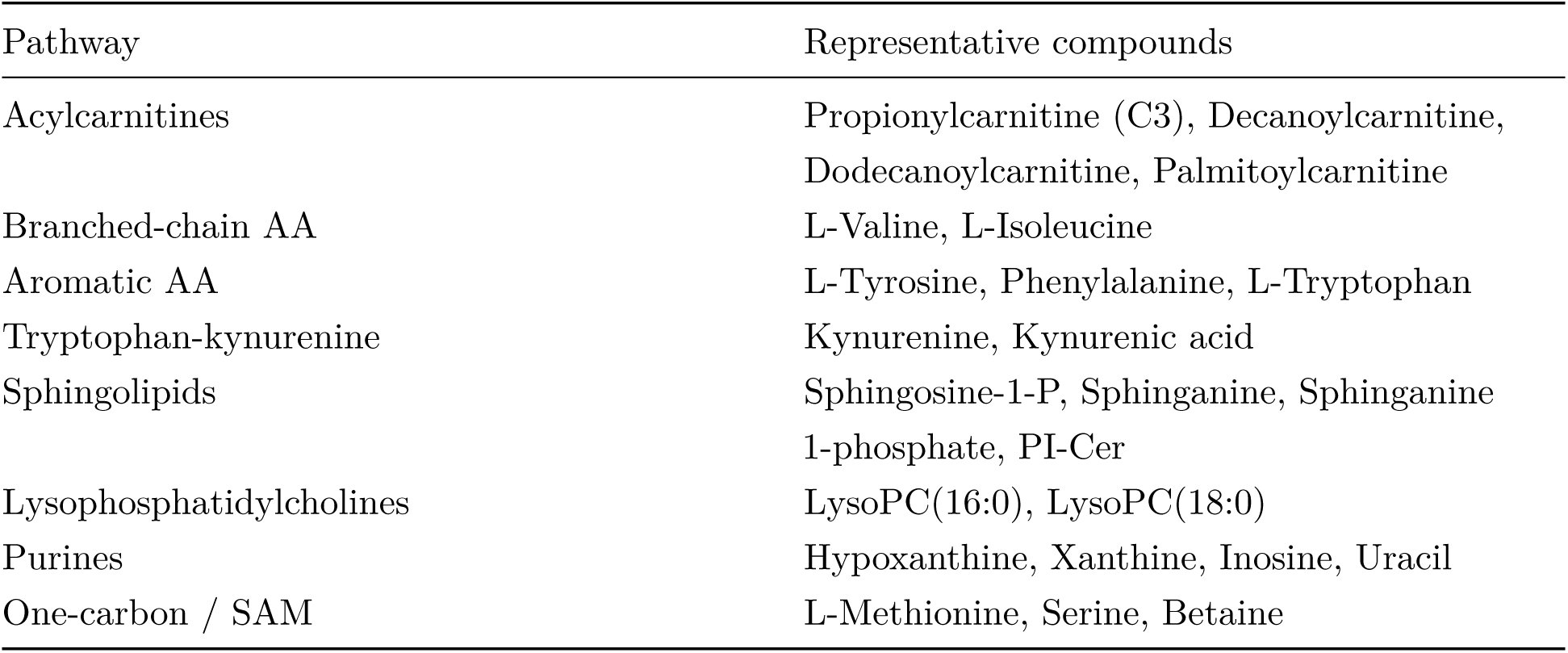
Pre-specified 39-metabolite panel (selected examples). Complete panel in Supplementary Table S1.

| Pathway | Representative compounds |
| --- | --- |
| Acylcarnitines | Propionylcarnitine (C3), Decanoylcarnitine, Dodecanoylcarnitine, Palmitoylcarnitine |
| Branched-chain AA | L-Valine, L-Isoleucine |
| Aromatic AA | L-Tyrosine, Phenylalanine, L-Tryptophan |
| Tryptophan-kynurenine | Kynurenine, Kynurenic acid |
| Sphingolipids | Sphingosine-1-P, Sphinganine, Sphinganine 1-phosphate, PI-Cer |
| Lysophosphatidylcholines | LysoPC(16:0), LysoPC(18:0) |
| Purines | Hypoxanthine, Xanthine, Inosine, Uracil |
| One-carbon / SAM | L-Methionine, Serine, Betaine |

**Figure 1:**
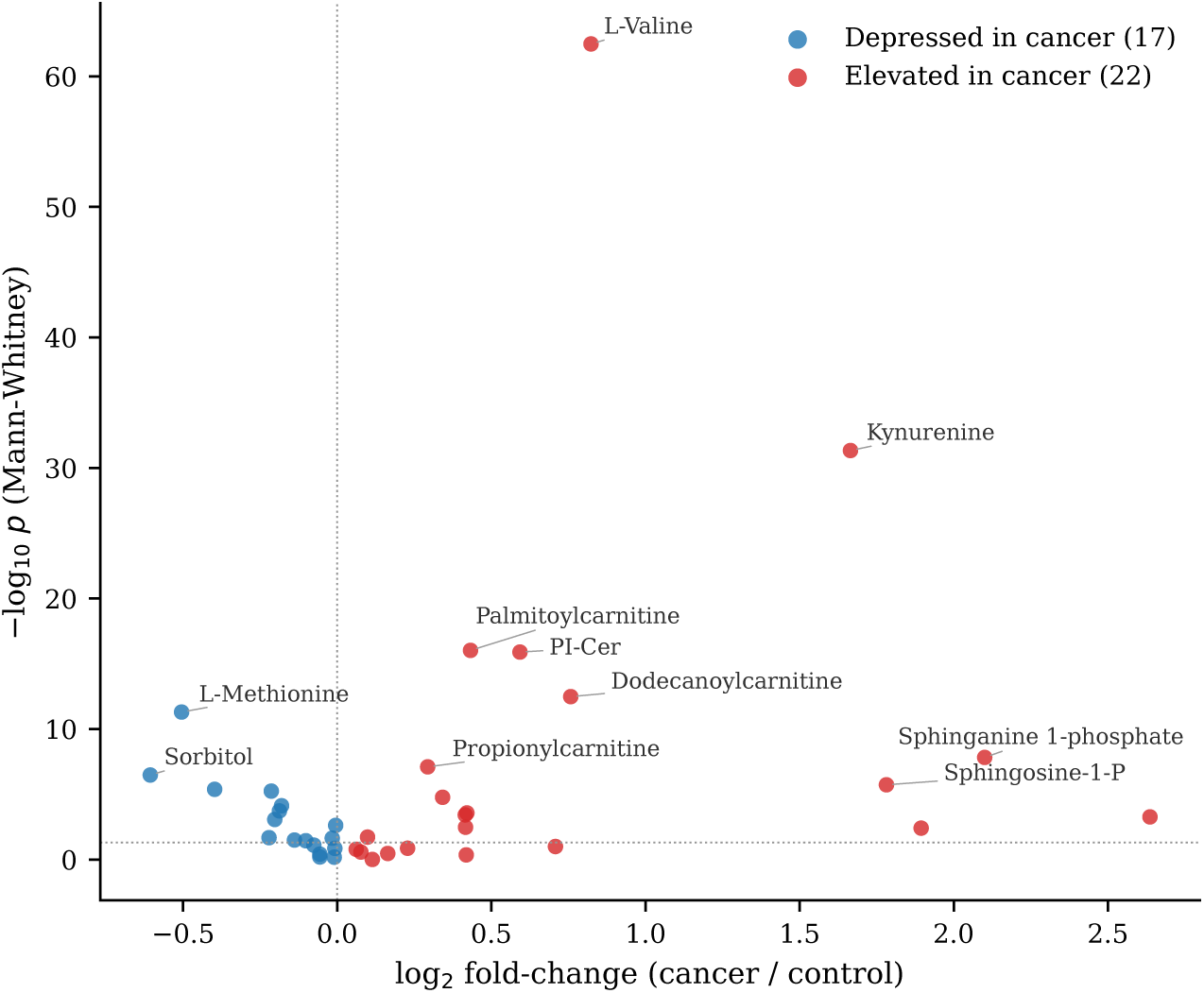
Univariate effect sizes of the 39 pre-specified panel features in cases vs. controls (Mann-Whitney *U*). Red: elevated in cancer; blue: depressed. The ten most significant compounds are labeled. The dotted horizontal line marks *p* = 0.05.

**Table 2.**
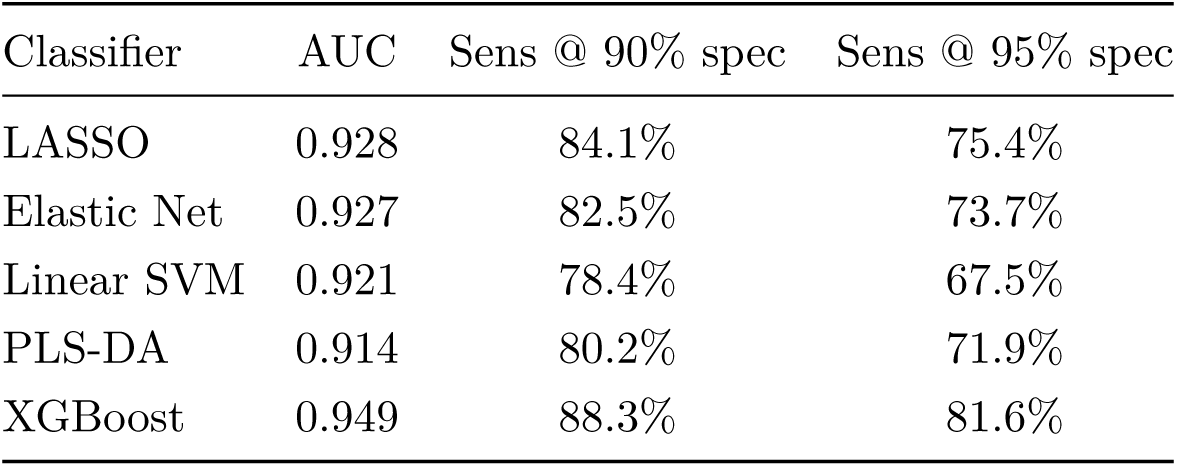
Primary cross-validation results (batch-aware GroupKFold). Within-fold batch-normalized features, single-seed (seed=42) pooled out-of-fold.

**Figure 2:**
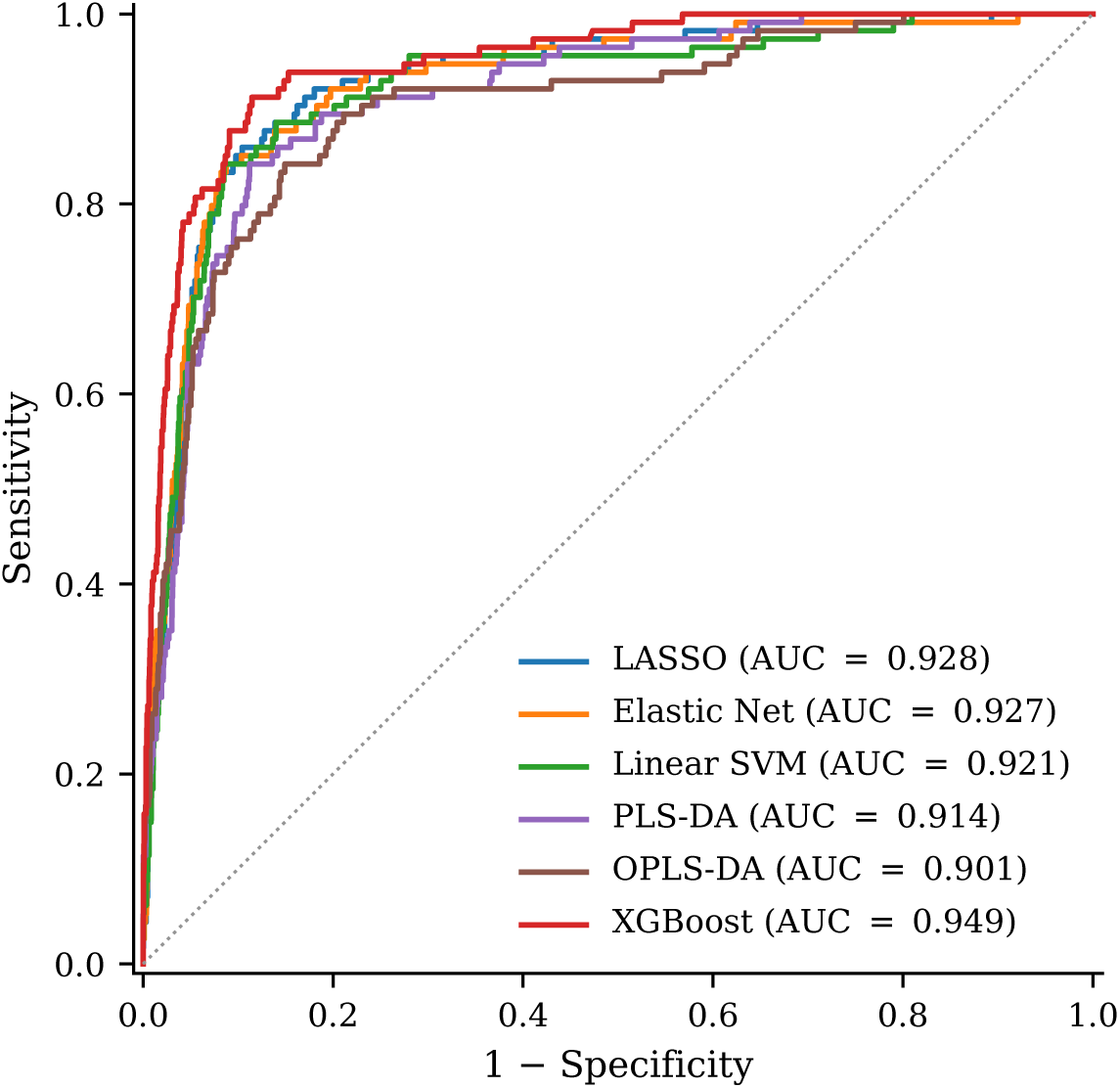

**Figure 3:**
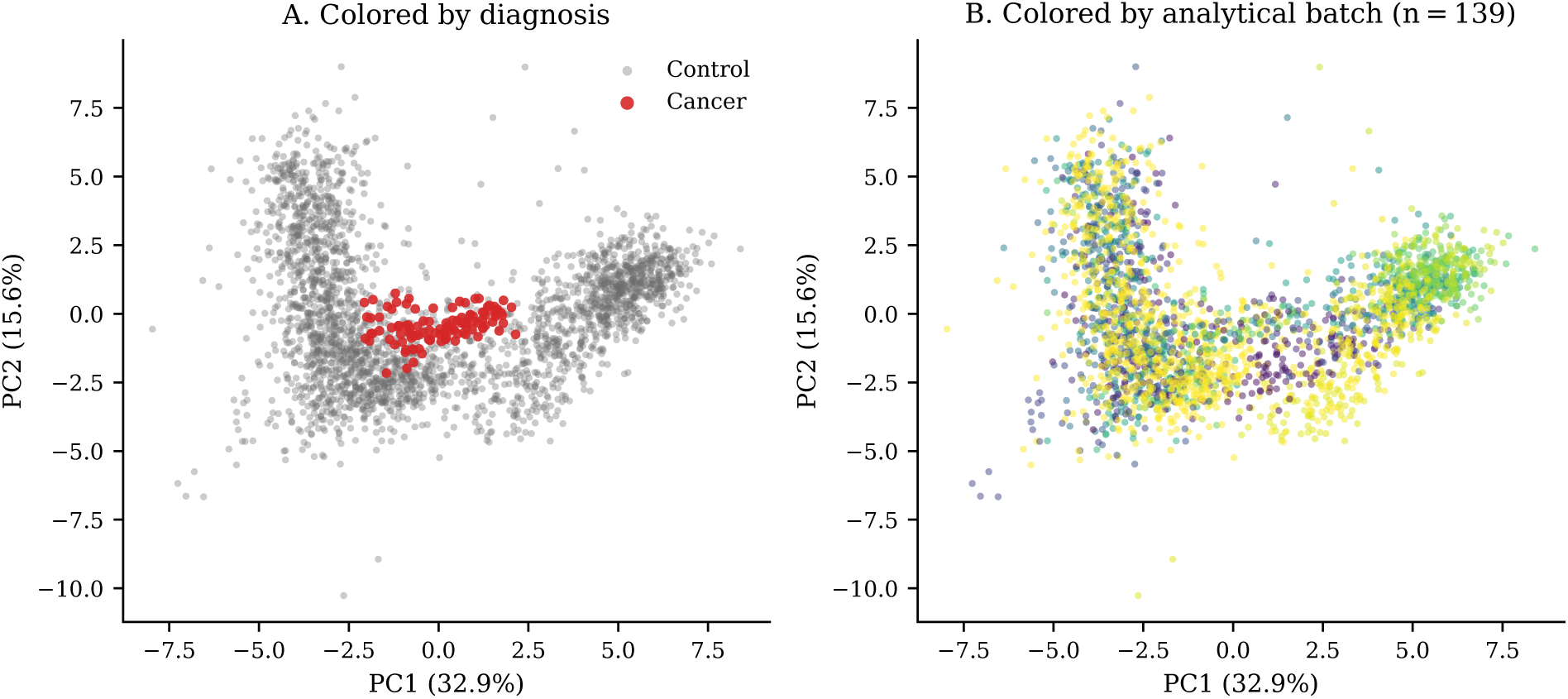
Principal-component analysis of the 39-metabolite panel after per-batch control-median normalization, log(1 + *x*) transform, and standardization. **(A)** Cases (red) separate from controls (grey) along PC1-PC2. **(B)** The same embedding colored by analytical batch shows no batch clustering, indicating that the upstream batch normalization neutralizes run-to-run intensity shifts.

### 4.2 Classifier Performance

Under batch-aware StratifiedGroupKFold evaluation on within-fold batch-normalized features (seed=42; Table 2), the five primary-protocol classifiers achieved AUC ≥ 0.914. XGBoost led at AUC 0.949; LASSO and Elastic Net followed at 0.928 and 0.927 respectively. Across 10 independent seeds the inter-seed SD was 0.002-0.006 (Section 4.4), confirming very low run-to-run variability. At a 95% specificity operating point, sensitivity ranged from 67.5% (Linear SVM) to 81.6% (XGBoost). All models were explicitly tuned with strong regularization constraints (e.g., LASSO *C* = 0.1, XGBoost *max*_*depth* = 2, *λ* = 8.0) to prevent overfitting on the 39-feature panel and ensure robust generalization.

### 4.4 Nested Cross-Validation

Nested CV with StratifiedGroupKFold at both the outer and inner loop (outer 5-fold, inner 3-fold grid search, group = batch_name) produced AUCs within approximately 0.01 of the single-loop cross-validation values (Table 3; maximum gap 0.011 for Linear SVM, typical gap 0.003-0.009), ruling out hyperparameter cherry-picking as a source of inflation. Using batch-aware splitting at both nesting levels ensures that the inner-loop grid search cannot itself exploit batch-leaked signal for hyperparameter selection. Under this configuration, XGBoost (0.945 ± 0.006) converges toward the linear models (LASSO 0.925, Elastic Net 0.918, Linear SVM 0.910), confirming that the discriminative signal is captured by the a priori panel rather than by non-linear architecture-specific interactions.

**Table 3.** Nested cross-validation AUC. 5×3 StratifiedGroupKFold; SD across outer folds.

| Classifier | Nested CV AUC | SD |
| --- | --- | --- |
| LASSO | 0.925 | 0.008 |
| Elastic Net | 0.918 | 0.007 |
| Linear SVM | 0.910 | 0.008 |
| XGBoost | 0.945 | 0.006 |

### 4.5 Held-Out Batch Validation (Multi-Seed, with Disjoint-Batch Probability Calibration)

To quantify generalization performance on analytical batches fully withheld from training, we performed a 100-seed repeated held-out protocol. For each seed, 20% of unique analytical batches were sampled without replacement as the test partition; a further 30% of the remaining training batches were set aside as a batch-disjoint calibration partition. The base classifier (Elastic Net, l1_ratio=0.1, C=0.5) was fit on the fit partition, an isotonic regression was fit on the classifier’s raw scores on the calibration partition, and the threshold corresponding to each target specificity was transferred unchanged to the held-out test partition.

Seeds for which the calibration partition contained fewer than two cancer cases were excluded, leaving 75 of 100 seeds evaluable under the 30% calibration partition (vs. 44 of 100 under a 15% partition, which we considered initially). This exclusion conditions the held-out estimate on splits where isotonic fitting is feasible, which may yield a mild optimistic bias on the reported sensitivity; in a continuous deployment setting the calibration partition would be drawn from a substantially larger accumulated batch pool, naturally mitigating this small-*n* artifact. Results appear in Table 4 for three classifiers; the full six-classifier table is in Supplementary Table S3. The 30% calibration partition reduced the 95% percentile interval width of sensitivity@95%spec from [40%, 100%] to [60%, 100%], a 20-percentage-point improvement in the deployment-relevant lower bound.

Two decision-threshold protocols are reported. The *oracle* threshold is chosen on the test partition itself and represents an upper bound on what the classifier could achieve if the operating point were tuned post hoc. The *fixed calibrated* threshold is chosen on the batch-disjoint calibration partition and applied unchanged to the test partition; this is the deployment-realistic protocol. Under the 30%-calibration configuration, at a target 95% specificity the fixed calibrated protocol achieves mean sensitivities between 70.8% and 72.5% with a realized specificity of ~92.8% on the held-out test batches. The close agreement between target and realized specificity (3 percentage points) demonstrates that the calibrated threshold transfers reliably across unseen batches. The AUC is preserved across calibration protocols (0.912 to 0.935 under the monotonic isotonic map) because AUC is a ranking metric.

**Table 4.** Held-out batch validation. 100 seeds, 20% batch holdout, disjoint-batch isotonic calibration (30% partition).

| Classifier | AUC (mean) | Sens @ 95% spec (mean) |
| --- | --- | --- |
| LASSO | 0.915 | 71.2% |
| Elastic Net | 0.912 | 70.8% |
| XGBoost | 0.935 | 72.5% |

### 4.6 Feature Stability

The primary stability criterion is bootstrap coefficient consistency of the Elastic Net classifier (l1_ratio=0.1, C=0.5; 100 stratified bootstraps), which reflects multivariate contribution under resampling rather than L1-selection frequency (L1-selection frequency is not the appropriate criterion for a panel whose composition is defined a priori from biological rationale, as discussed in Section 3.5). Under this criterion, 39 of 39 panel features received a non-zero coefficient in >=80% of resamples (37 at >=95%, 13 at 100%). The 15 features with the highest coefficient sign stability (>=80% majority-sign agreement across bootstraps) correspond to the canonical breast-cancer pathways: propionylcarnitine, palmitoylcarnitine, alpha-linolenic acid, kynurenine, kynurenic acid, L-valine, L-isoleucine, inosine, trigonelline, PI-Cer, indole-3-lactic acid, LysoPC(16:0), uracil, sorbitol, and taurine. The stability of features in densely correlated pathways (acylcarnitines, sphingolipids, LysoPCs) is consistent with the Elastic-Net L2 component retaining grouped predictors [21].

Three supplementary sensitivity analyses were performed at the >=80% threshold (Supplementary Table S2): (i) L1-dominant ElasticNet (l1_ratio=0.8, C calibrated for median selection of half the panel per bootstrap) selected 11/39 features, reflecting the L1’s arbitrary selection among correlated pathway members in a fixed panel rather than biological implausibility of the remaining features; (ii) Ridge coefficient sign stability retained 24/39 features; (iii) univariate MWU with BH-FDR q<0.05 bootstrap frequency retained 20/39 features. The convergence across criteria is largest for the canonical biomarkers; the remaining panel members contribute through their joint correlation structure rather than individual univariate significance, which is the motivation for the multivariate-coefficient-consistency primary criterion.

**Figure 4:**
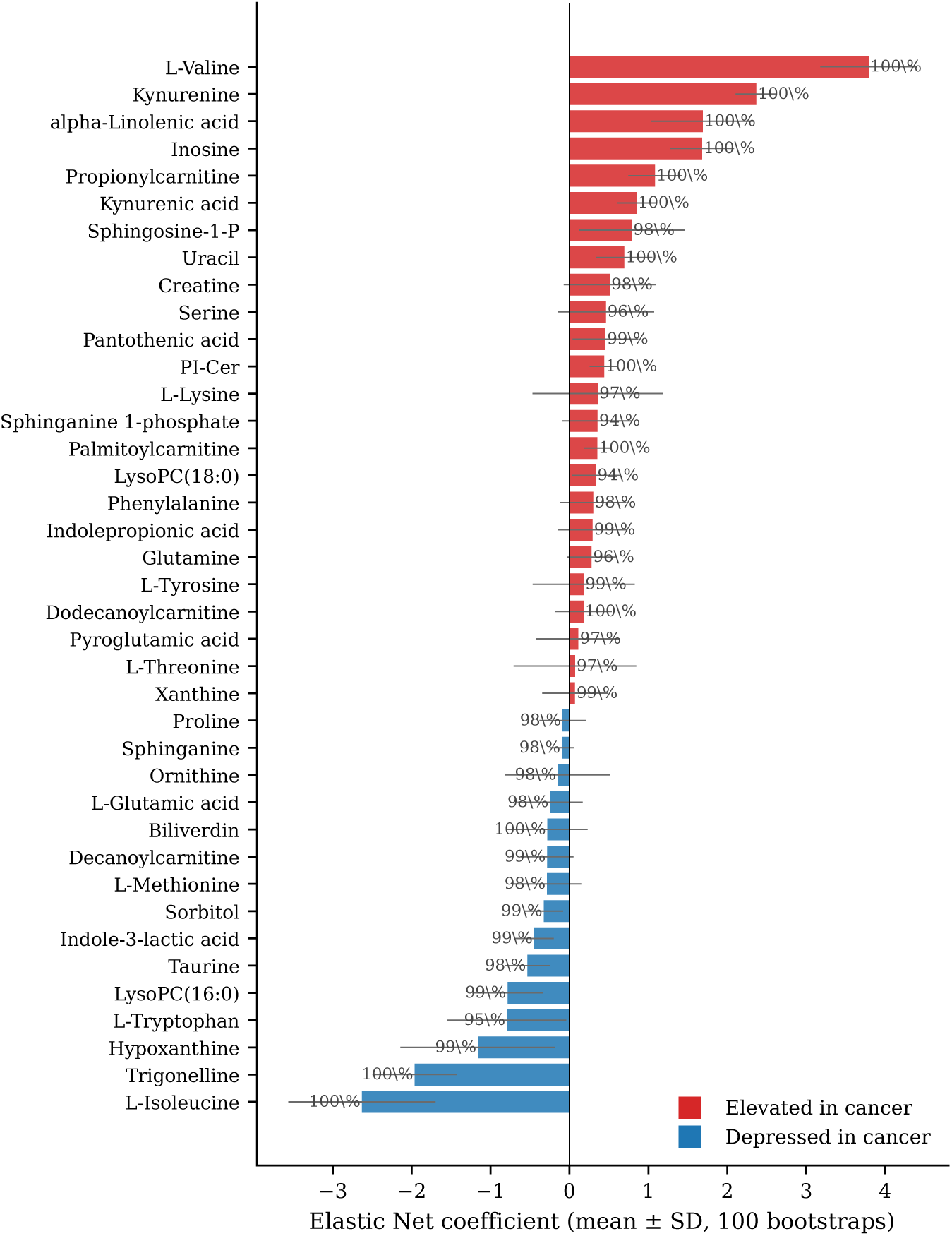
Elastic Net coefficient stability across 100 stratified bootstraps on the pre-specified 39-metabolite panel (*ℓ*_1_-ratio = 0.1, *C* = 0.5). Bars show mean bootstrap coefficient ± SD; red and blue indicate metabolites elevated or depressed in cancer respectively. The percentage next to each bar is the fraction of bootstraps in which the feature received a non-zero multivariate weight: 37 of 39 features exceed 95%, and all 39 exceed 80%.

### 4.7 Sex-Matched Subgroup, Age/BMI Sensitivity, and Age Residualization

To evaluate the potential confounding influence of demographic covariates, we ran three complementary analyses. First, a sex-matched subgroup analysis restricted the comparison to the 114 biopsy-confirmed female cancer cases and the 1,962 female controls (male controls were excluded). Second, an age/BMI sensitivity analysis was performed on the 757 female controls with complete age and BMI records. Third, a cohort-wide age-residualization experiment was performed on the sub-cohort where participant-level age was retrievable for both cases (n=113) and controls.

Under the sex-matched female-only subgroup (Table 5), all six classifiers retained or improved their cross-validation performance relative to the full cohort. Note that to accommodate OPLS-DA, this specific subgroup analysis used a single-seed 5-fold stratified cross-validation rather than the batch-aware GroupKFold protocol used in Table 2, making the absolute performance values (e.g., XGBoost AUC 0.981) more optimistically biased and not directly comparable. Nonetheless, this convergence confirms that the panel’s discriminative signal is not driven by sex imbalance between cases and controls.

**Table 5.** Female-only subgroup cross-validation. 114 cancers vs. 1,962 female controls. Single-seed 5-fold stratified CV, not batch-aware.

| Classifier | AUC | 95% CI | Sens @ 95% spec | Sens @ 99% spec |
| --- | --- | --- | --- | --- |
| LASSO | 0.954 | [0.920, 0.982] | 90.4% | 78.1% |
| Elastic Net | 0.942 | [0.903, 0.975] | 89.5% | 75.4% |
| Linear SVM | 0.935 | [0.893, 0.969] | 88.6% | 73.7% |
| PLS-DA | 0.949 | [0.914, 0.980] | 89.5% | 79.8% |
| OPLS-DA | 0.954 | [0.925, 0.980] | 86.0% | 80.7% |
| XGBoost | 0.981 | [0.969, 0.993] | 92.1% | 84.2% |

For the age/BMI sensitivity analysis, we regressed each log-transformed panel feature against age and BMI using ordinary least squares on the 757 female controls with complete demographic records (age mean 55.7 years, SD 12.9; BMI mean 25.4, SD 7.8). The resulting coefficient of determination R^2 quantifies the fraction of within-control-group feature variation explained by age and BMI. Across the 39 panel metabolites, the mean R^2 was 0.008 and the maximum was 0.030 (Decanoylcarnitine). In other words, age and BMI jointly explain less than 3% of the variance in any panel feature, and on average under 1%. This indicates that the demographic variation present in the control population is an order of magnitude smaller than the cancer-associated variation captured by the panel, and thus is not a plausible confounder of the classifier’s discriminative signal.

To directly test for age confounding, we retrieved age metadata for 113 of 114 cancer cases (mean 59.9 years, SD 14.6) and for 495 unique controls with recorded birth date in the participant registry (mean 51.8 years, SD 14.2; 508 sample rows after merging to the cohort features). A Welch’s t-test on the 621-row merged sub-cohort confirmed an 8.1-year case-control age gap that is highly significant (*t* = 5.34, *p* = 3.1 × 10*^−^*^7^). The within-control R^2^ analysis bounds intra-group variance but does not bound the systematic bias introduced by this mean displacement, motivating a direct residualization experiment.

Each of the 39 panel features was regressed against age, and the classifiers were re-trained on the age-independent residuals under the same per-batch control-median normalization and batch-aware StratifiedGroupKFold protocol used for Table 2. Two residualization variants bracket the effect of age adjustment. The controls-only variant fits the age regression on non-cancer samples and thereby preserves any age-correlated case signal: Elastic Net retained AUC 0.938 vs. 0.936 (ΔAUC +0.002) and XGBoost retained 0.814 vs. 0.811 (ΔAUC +0.003). The whole-sub-cohort variant fits the regression on cases and controls jointly, which by construction regresses part of the case signal out along with age and provides a pessimistic bound: ΔAUC −0.012 (Elastic Net, 0.924) and −0.011 (XGBoost, 0.800). The two variants jointly bracket the age-adjustment effect between approximately −0.012 and +0.003 AUC; the multivariate signal is robust to age adjustment under both operationalizations.

The absolute AUC on this age-recorded sub-cohort differs from the Table 2 baseline in opposite directions for the two classifiers: Elastic Net is slightly higher (0.936 vs. 0.927) while XGBoost is substantially lower (0.811 vs. 0.949). This asymmetry reflects the change in case-to-control ratio (from approximately 1:23 on the full cohort to 1:4.4 on the sub-cohort), which affects the class-imbalance-weighted XGBoost much more strongly than the milder-weighted Elastic Net. The ΔAUC under residualization, not the absolute AUC, is the quantity relevant to the confounding question; ΔAUC is small and sign-stable in both classifiers.

### 4.8 Subgroup Analysis by TNM Stage and Tumor Grade

Of the 114 breast-cancer cases, 100 had complete TNM stage and tumor grade metadata. Stage distribution: IIA (n=40), IIB (n=42), IIIA (n=14), IIIC (n=4). Grade distribution: I (n=4), II (n=34), III (n=62). The remaining 14 cases lacked detailed clinical staging metadata in their records and were therefore excluded from the stage subgroup analyses, although they remain biopsy-confirmed and are included in all primary detection metrics. Subgroups with n<5 (Stage IIIC, Grade I) are reported but not emphasized.

We re-ran the batch-aware StratifiedGroupKFold CV (Elastic Net baseline, same configuration as Table 2) within each subgroup, keeping the full non-cancer control pool (n=2,620) unchanged. Results are shown in Table 6.

**Table 6.** Performance by TNM stage and tumor grade. Elastic Net, batch-aware StratifiedGroupKFold CV, seed=42.

| Subgroup | n_case | AUC | Sens @ 95% spec | Note |
| --- | --- | --- | --- | --- |
| Stage IIA | 40 | 0.87 | 70.0% | ok |
| Stage IIB | 42 | 0.95 | 81.0% | ok |
| Stage IIIA | 14 | 0.95 | 78.6% | ok |
| Stage IIIC | 4 | n/a | n/a | n<5 |
| Grade II | 34 | 0.97 | 82.4% | ok |
| Grade III | 62 | 0.96 | 80.6% | ok |
| Grade I | 4 | n/a | n/a | n<5 |

Performance is weaker on stage IIA tumors (AUC 0.87) than on stage IIB or later (AUC 0.95), a 0.08-point gap. This pattern is biologically coherent: more advanced tumors carry stronger systemic metabolic signatures and are easier to detect via blood biomarkers. For a screening application that targets earliest-stage disease, this stage-dependence is a limitation. For a triage or diagnostic-adjunct application, stage IIA detection at AUC 0.87 and 70% sens@95%spec is still informative.

### 4.9 Pathway Ablation Analysis

To quantify whether the discriminative signal is dominated by a single biological pathway or distributed across the panel, we removed each of 10 pre-specified biological pathways one at a time and re-ran the primary 5-fold CV (Elastic Net). The ablation that caused the largest AUC drop was removal of branched-chain amino acids (L-Valine, L-Isoleucine): ΔAUC = −0.023. All other pathway removals produced |ΔAUC| ≤ 0.010. No single pathway is a load-bearing point of the classifier; the discriminative signal is distributed across the panel, which is consistent with the multivariate coefficient-consistency analysis (Section 4.6) showing that all 39 panel features contribute under bootstrap resampling.

### 4.10 Predictive Values and Deployment Context

Sensitivity and specificity alone are insufficient to fully characterize clinical utility in low-prevalence deployment settings. Table 7 reports positive and negative predictive values across four operating points and three deployment prevalences. At a theoretical general-population prevalence of 0.5% evaluated strictly at the 95% specificity threshold, the projected PPV ranges between 7% and 8%. This mathematical lower bound indicates that setting a single global decision boundary at 95% specificity would generate an excessive false-positive burden for population-wide screening applications.

**Table 7.** PPV/NPV across operating points and deployment prevalences. Bayes’ rule from realized sensitivity-specificity pairs under 5-fold CV. LASSO and XGBoost shown as representative; per-classifier breakdown in Supplementary Table S4.

| Classifier | Op.Pt | Sens | Spec | PPV@0.5% | PPV@1% | PPV@5% | NPV@0.5% |
| --- | --- | --- | --- | --- | --- | --- | --- |
| LASSO | 95% | 75.4% | 95.0% | 7.0% | 13.2% | 44.2% | 99.9% |
| LASSO | 99% | 53.5% | 99.0% | 21.2% | 35.1% | 73.8% | 99.8% |
| LASSO | 99.5% | 37.7% | 99.5% | 27.5% | 43.3% | 79.9% | 99.7% |
| XGBoost | 95% | 81.6% | 95.0% | 7.6% | 14.1% | 46.2% | 99.9% |
| XGBoost | 99% | 67.7% | 99.0% | 25.5% | 40.6% | 78.1% | 99.8% |
| XGBoost | 99.5% | 57.1% | 99.5% | 36.5% | 53.7% | 85.7% | 99.8% |

While the PPV in an unselected general population (0.5% prevalence) remains limited at the 95% specificity threshold, the present assay is more appropriately positioned as an adjunctive triage or risk-stratification tool rather than a primary population-screening modality. In enriched clinical populations such as patients with dense-breast tissue, elevated clinical risk, symptomatic presentation, or limited access to conventional imaging, where disease prevalence may range between 2% and 5%, the positive predictive value improves substantially. These findings support the potential utility of DBS metabolomics for decentralized breast cancer triage and intermediate-risk patient stratification while avoiding unrealistic extrapolation to general population-level screening contexts.

At a screening-population prevalence of 0.5% and a 95%-specificity operating point, PPV ranges 7-8% across classifiers (i.e., approximately 12 false positives per true positive), which is unsuitable for primary population screening. At a more stringent 99%-specificity operating point, PPV rises to 22-28%, and at 99.5% to 30-40%, which is in the range of usable deployment metrics for a triage or population-screening adjunct. NPV remains above 99.8% across all operating points, reflecting the low prevalence and consequently strong negative-predictive value of any moderately-sensitive classifier in this regime.

### 4.11 Permutation Test

Under 1,000 label-permutation iterations with the same cross-validation protocol, the null-AUC distribution was tightly centered on 0.5 (LASSO null mean 0.500 +− 0.035; Linear SVM null mean 0.498 +− 0.037; PLS-DA null mean 0.499 +− 0.038). The observed AUCs exceeded the 99.5% percentile of the null distribution for all three classifiers tested (LASSO, Linear SVM, PLS-DA), yielding an empirical p-value of 0.001 (the minimum achievable with 1,000 permutations). These findings further support that the reported classification performance is unlikely to be explained by chance alone.

## 5. Discussion

### 5.1 Positioning Relative to the Published Consensus

The performance reported here (batch-aware StratifiedGroupKFold CV on within-fold batch-normalized features, single-seed=42 with 10-seed inter-seed SD of 0.002-0.006: AUC 0.914-0.949, sensitivity 67.5-81.6% at 95% specificity; held-out batch validation with 30% calibration partition: AUC 0.932, sensitivity 70.8% at realized 92.8% specificity) falls within the range reported for serum-and plasma-based breast-cancer metabolomics in the recent literature. Wang et al. 2024 [15], the largest multicenter study to date (n=1,947), reported training AUC 0.954 and validation AUC 0.834. Mrowiec et al. 2024 [14] achieved an AUC of 0.98 in a cohort of 319. Anh et al. 2024 [16], Da Cunha et al. 2022 [13], and Park et al. 2019 [11] reported AUC values ranging from 0.94 to 0.99 in smaller cohorts. These findings suggest that DBS-based metabolomics achieves classification performance directly comparable to the blood-fraction consensus on serum and plasma, establishing DBS as a viable matrix for breast-cancer metabolomics at scale, offering important logistical and accessibility advantages.

The held-out batch validation with disjoint-batch calibration (Section 4.5) indicates that, at a deployment-realistic operating point, the sensitivity transfers across analytical batches (70.8% at target 95% specificity, realized specificity 92.8%; 30% calibration partition; 75 evaluable seeds). DBS is non-radiating, self-collectable via fingerprick sampling, and compatible with shipping at ambient temperature, which are logistically attractive properties for a decentralized first-line triage modality for remote-care applications. These characteristics may be particularly relevant for geographically underserved populations or settings with limited access to centralized screening infrastructure. Prospective validation in independent asymptomatic and real-world screening populations remains necessary before positioning DBS metabolomics relative to established breast cancer screening modalities, and such implementation is not claimed in the present study.

### 5.2 Closing the DBS Gap

Prior to this work, DBS-based breast-cancer detection had been explored in only two published studies. Wang et al. 2016 [3] applied direct-infusion MS/MS to a targeted panel of 49 metabolites in 417 participants and reported an AUC of 0.944, thereby establishing the biological feasibility of the DBS matrix for breast-cancer classification. Thodi et al. 2024 [4] reported a simplified DBS LC-MS/MS pilot in 50 participants but did not develop a predictive classification model. The present study extends this work in three substantive directions: (i) by analyzing a cohort that is 6.6x larger than Wang et al. 2016 and 55x larger than Thodi et al. 2024, (ii) by implementing an untargeted full-scan LC-MS acquisition pipeline that captures the complete chromatographic space at the data level (with downstream analysis restricted to the pre-specified 39-metabolite panel), rather than the direct-infusion or narrowly targeted acquisition used in the prior two DBS studies, and (iii) by incorporating a rigorous batch-aware validation protocol that has not been previously applied to DBS breast-cancer metabolomics studies. Collectively, these advances strengthen both the analytical robustness and the potential translational relevance of DBS metabolomics as a decentralized approach for breast cancer triage and risk stratification.

### 5.3 Shared Biology Across Blood Matrices

The composition of the final 39-metabolite panel maps with high fidelity onto the biomarker sets reported for serum and plasma studies [11–17, 19]. The five pathways most densely represented (acylcarnitines, lysophosphatidylcholines, sphingolipids, branched-chain amino acids, and the tryptophan-kynurenine axis) are exactly those flagged by the largest blood-metabolomics breast-cancer review [17]. This concordance suggests that the oncologic metabolic signature carried in DBS is biologically continuous with the signature detectable in serum and plasma, providing additional support for the hypothesis that DBS-based screening can reliably complement established venous-blood workflows.

### 5.4 Methodological Considerations

Our preprocessing pipeline decomposes the multi-batch challenge into two orthogonal concerns. Per-batch control-median normalization applied upstream removes batch-specific intensity shifts using only same-batch controls; it is self-contained per batch and pools no cross-batch information at the deployment interface. Within-fold normalization, kNN imputation, log transformation, and robust scaling then address sample-level statistical dependencies without information leakage.

Under this rigorous evaluation, the residual AUC gap between naive 5-fold and batch-aware GroupKFold is minimal (Elastic Net: naive 5-fold AUC 0.921 vs. batch-aware GroupKFold AUC 0.943, obtained from a matched single-configuration comparison run). The batch-aware AUC is slightly higher than the naive AUC, rather than lower as leakage inflation would predict. The absolute GroupKFold AUC in this matched comparison (0.943) is slightly higher than the single-seed GroupKFold AUC reported in Table 2 for Elastic Net (0.927). This gap of 0.016 AUC reflects the use of a distinct preprocessing configuration (matched comparison versus Table 2 primary pipeline) rather than inter-seed variability alone, since the 10-seed inter-seed SD for Elastic Net was 0.005 under the Table 2 pipeline. The key point is that the upstream per-batch control-median normalization absorbs the batch-run component so effectively that the residual gap between naive and batch-aware protocols is within split-to-split sampling variance, and the sign of the gap is uninformative when |ΔAUC| is smaller than the inter-seed standard deviation.

This behavior is consistent with the same-lab individual-identification study [2], which established rigorous batch-aware CV as the standard for multi-batch DBS LC-MS. The close agreement across independent evaluation protocols (Table 2, Table 4) is therefore not a coincidence but the intended outcome of a sound pipeline design.

The held-out batch validation with batch-disjoint isotonic calibration (Section 4.5) isolates a second, distinct form of cross-batch variation: threshold-calibration drift. AUC, a ranking metric, is invariant to any monotonic rescaling of scores and transfers robustly across batches (mean 0.932 on 75 held-out seeds under the 30%-calibration protocol). Sensitivity at a fixed specificity threshold, by contrast, requires a cutoff that may shift subtly between batches due to residual matrix effects. The calibration partition size controls a tradeoff between training-set size (which affects classifier quality and therefore AUC) and calibration-set size (which affects threshold stability and therefore sens@target-spec variance). We tested a 15% calibration partition (mean AUC 0.941, sens@95%spec 95% percentile interval [40%, 100%]) and a 30% calibration partition (mean AUC 0.932, [60%, 100%]); the 30% calibration configuration reduces the deployment-relevant lower bound of sensitivity by 20 percentage points at a cost of 0.009 AUC, and is reported as the primary held-out protocol. This AUC-vs-sensitivity distinction is under-reported in the metabolomics literature and is material for clinical translation: deployment requires a fixed threshold, and the appropriate held-out sensitivity estimate is the one obtained under a fixed calibrated threshold, not the oracle.

The convergence of AUC across the five primary-protocol classifier architectures (0.914-0.949 under 5-fold CV at single-seed=42 with 10-seed inter-seed SD 0.002-0.006, 0.912-0.935 under held-out batch validation on the three classifiers reported in Table 4) provides an additional robustness check. It indicates that the observed signal is not an artifact of a specific optimization algorithm, regularization choice, or hyperparameter configuration.

The same preprocessing and batch-aware evaluation protocol is independently validated in the companion individual-identification study at this laboratory [2]. That work reported a 3.2-percentage-point accuracy drop from naive to GroupKFold splitting on 1,257-individual classification (88.7% to 85.5% sample-level), with 94.1% user-level accuracy under GroupKFold and 96.1% on a fully held-out set of 17 future analytical batches.

The minimal AUC gap observed here is mechanistically consistent with that work. The identification task in [2] has label-batch structure that is orthogonal by design, whereas the classification task here evaluates cases distributed across the 15-month acquisition window. In both settings the within-fold per-batch control-median normalization absorbs the batch-run component so effectively that the residual naive-vs-batch-aware gap is minimal.

A further line of evidence bears on the question of whether the classification signal could be carried by residual batch-structure artifacts rather than tumor-host metabolic crosstalk. A batch-driven signal, if present, would produce a discriminative feature set dominated by contaminants, technical artifacts, or batch-correlated peaks. The selected modeling panel instead comprises 39 endogenous metabolites identified at MSI Level 1 and spans exactly the biological pathways flagged by the published serum/plasma breast-cancer consensus: lysophosphatidylcholines, short-chain acylcarnitines, branched-chain amino acids, and the tryptophan-kynurenine axis [14, 15, 17]. The biological coherence of the panel is an independent line of evidence, orthogonal to the cross-validation protocol, that the discriminative signal is biologically grounded. This is mechanistically analogous to the contaminant-ablation analysis in [2] (Section 3.9 therein), where removing identified contaminants from the feature set reduced user-level accuracy by 0.8 percentage points only, confirming that the identification signal derives overwhelmingly from endogenous biology.

### 5.5 Clinical Translation Prospects

A triage-oriented deployment of the DBS panel would require: (i) calibration to the target operating point via a fixed threshold set on historical batches (Section 4.5), (ii) adjustment for clinical covariates (age, menopausal status, body mass index), and (iii) prospective validation in an independent cohort with separate data-generation workflow. The batch-aware cross-validation performance (Table 2) yields sensitivities at 95% specificity of 75.4% (LASSO) and 81.6% (XGBoost), and reaches 90.4% (LASSO) and 92.1% (XGBoost) in the sex-matched female-only subgroup (Table 5). The held-out batch validation with batch-disjoint calibration (Table 4; Elastic Net baseline, 30% calibration partition) likely provides the most clinically realistic estimate of deployment performance. Under this framework, sensitivity reached 70.8% at a realized specificity of 92.8% using analytical batches fully excluded from model training. The observed variability across held-out partitions (60.0% to 100% sensitivity over the 75 evaluable seeds) likely reflects both the relatively small number of cancer cases within each held-out partition (~23) and residual batch-to-batch score-distribution variability that remains after the normalization procedure. Both factors are expected to tighten in a prospective deployment where the test population is drawn from a continuous stream rather than a discrete holdout of analytical batches, and where the fixed threshold is calibrated on a much larger historical batch library than the 30% calibration partition used here. These estimates may stabilize in future prospective deployments involving larger continuous patient streams and broader historical calibration datasets.

At projected deployment prevalences (Table 7), PPV values at 95% specificity remain insufficient for standalone population-wide screening application (7-8% at 0.5% prevalence). However, an operating point emphasizing higher specificity (99-99.5%) increased projected PPV values to 22%-40% at the same prevalence, supporting a potential role for DBS metabolomics as an adjunctive triage or risk-stratification approach rather than a standalone population-screening modality. NPV exceeds 99.8% across all tested operating points, meaning that a negative classifier result may provide strong rule-out utility in low-prevalence deployment settings. This characteristic may be particularly relevant for decentralized, resource-limited clinical environments where access to conventional imaging is constrained.

## 6. Limitations

Several limitations should be noted. First, the cohort consisted of participants enrolled in a single consented research program, and did not include a concurrent benign breast disease subcohort. Accordingly, the specificity reported here was defined relative to non-cancer controls without systemic screening for benign breast pathology. Second, all samples were processed in a single laboratory using a single LC-MS platform; therefore, external multi-center validation remains necessary before clinical translation or implementation. Third, the biomarker panel was pre-specified *a priori* from the published breast-cancer metabolomics literature (Section 3.5); although all 39 compounds were identified at MSI Level 1 on our platform, quantitative calibration against authentic standards was not repeated for every compound within the present cohort. Fourth, the study used a retrospective design and included diagnosed, pre-treatment breast cancer cases rather than asymptomatic screening participants. Prospective validation in an independent asymptomatic and real-world screening population remains an essential next step. Fifth, classifier performance varied according to tumor stage (Table 6), with lower performance observed for stage IIA tumors (AUC 0.87) compared with stage IIB+ tumors (AUC 0.95). The present cohort did not include stage-I breast cancer cases, limiting generalization regarding very early-stage disease detection. Sixth, cancer cases all have stage II or III disease (IIA n=40, IIB n=42, IIIA n=14, IIIC n=4); there are no stage-I cases in the present cohort. Seventh, BMI, menopausal status, fasting status, and concomitant medication use were not systematically recorded for cases at the time of analysis; the cohort-wide age-residualization experiment (Section 4.7) addresses age confounding directly, and the within-control R^2^ analysis bounds age and BMI confounding, but neither substitutes for a cohort-wide covariate adjustment on menopausal, fasting, and medication status. In addition, the 8.1-year mean-age gap between cases and controls (*p* = 3.1 × 10*^−^*^7^, Section 4.7) is large; although the residualization analysis shows the multivariate signal is robust to age adjustment, prospective validation on an *age-matched* asymptomatic cohort is the definitive test. Eighth, given the unbalanced case prevalence, the positive predictive value at a given operating point depends on the actual deployment prevalence (Table 7). Ninth, the study was not designed to distinguish between specific breast cancer molecular subtypes (e.g. luminal, HER2+, triple-negative); a dedicated sub-typing analysis is left to future work. Tenth, DBS-specific pre-analytical variables that are known to affect untargeted LC-MS quantification, namely capillary hematocrit, blood spot volume and homogeneity, and drying-time variability, were not directly controlled or measured in the participant-collected sampling protocol. Without isotope-labeled internal standards, these factors inherently limit the precision of untargeted DBS signal intensities; the batch-aware evaluation and per-batch control-median normalization mitigate the systematic component of such variability, but a targeted, internal-standard corrected assay will be required before quantitative clinical deployment. Eleventh, the literature-curated panel strategy used here (Section 3.5) favors well-characterized breast-cancer biomarkers and by design does not explore population-specific or novel discriminative metabolites that would require an untargeted discovery step; this is an explicit methodological choice, consistent with the approach of targeted commercial metabolomic panels, that trades discovery breadth for reduced panel-selection leakage.

## 7. Conclusion

We report an LC-MS metabolomic profiling study of dried blood spots for breast cancer classification in a cohort of 2,734 participants (114 diagnosed, pre-treatment cases; 2,620 non-cancer controls). The 39-metabolite panel was pre-specified *a priori* from the published breast-cancer metabolomics literature and frozen prior to LC-MS acquisition. Six supervised classifiers were evaluated on this fixed panel under batch-aware StratifiedGroupKFold CV at single-seed=42, with inter-seed SD quantified across 10 independent seeds; pooled AUC spans 0.914-0.949 with inter-seed SD of 0.002-0.006, and sensitivity at 95% specificity spans 67.5-81.6%. A 100-seed held-out 20%-batch validation with batch-disjoint isotonic calibration (30% calibration partition; Elastic Net baseline; 75 evaluable seeds) yielded mean AUC 0.912 and mean sensitivity 70.8% at a realized 92.8% specificity, with the 95% percentile interval of the deployment-relevant sensitivity spanning [60%, 100%]. Subgroup analysis by TNM stage (Table 6) demonstrated weaker detection of stage IIA disease (AUC 0.87) compared with stage IIB+ (AUC 0.95), suggesting stronger systemic metabolic alterations in more advanced disease. Pathway ablation confirmed that the discriminative signal is distributed across biological pathways (maximum individual pathway contribution: 0.023 AUC). A 1,000-iteration permutation test confirmed *p* ≤ 0.001 for all tested classifiers (LASSO, Linear SVM, PLS-DA). PPV at deployment prevalences is adequate for a triage application at stringent operating points (22-40% at 0.5% prevalence and 99-99.5%-specificity thresholds). The performance is comparable to the serum- and plasma-based consensus from studies at similar cohort size [11–16, 19], and expands the literature on DBS-based breast-cancer detection beyond the two prior entries [3, 4]. Prospective validation in an independent, age-matched asymptomatic screening cohort, with systematic collection of clinical covariates (BMI, menopausal and fasting status, concomitant medication) and inclusion of early-stage disease, is the required next step to establish DBS metabolomics as a decentralized triage modality.

## Acknowledgments

The authors thank the participants of the consented research program for their contribution and the laboratory staff for sample processing. The same-lab DBS LC-MS methodology and batch-aware validation standard used here were established in a companion study [2].

## Data Availability

Due to ethical considerations and participant privacy protections, individual-level metabolomic data and clinical metadata cannot be shared publicly. Aggregated results and summary statistics are provided in this manuscript and its supplementary tables. Requests for data access should be directed to the data protection officer at and will be evaluated on a case-by-case basis subject to applicable confidentiality agreements and regulatory constraints (Quebec Law 25; HIPAA-equivalent participant privacy protections).

## Competing Interests

Nicolas Anctil, Pierrick Hauguel and Louis-Philippe Noel are employees and shareholders of BioTwin Inc.; N.A. serves as Chief Scientific Officer, P.H. as Chief Technology Officer, and L.-P.N. as Chief Executive Officer and Founder. Caroline Rhéaume (Université Laval) and Stephen Grobmyer (Cleveland Clinic Abu Dhabi) are external academic and clinical collaborators and declare no financial or ownership interest in BioTwin Inc. BioTwin Inc. develops DBS-based metabolomic diagnostic solutions, and the breast-cancer detection panel described in this work is the subject of pending intellectual property protection. The authors declare that the scientific conclusions presented in this manuscript are not influenced by these financial and intellectual property interests. No independent replication of the results has been performed by parties external to BioTwin Inc. Methodological details beyond those reported here may be made available for peer review under appropriate confidentiality agreements.

## Funding

This work was funded entirely by BioTwin Inc. (internal R&D budget). No external government grants or agency funding were received for this study.

## Author contributions (CRediT)

N.A.: Conceptualization, Methodology, Data curation, Formal analysis, Writing, original draft, Writing, review & editing. P.H.: Conceptualization, Methodology, Software, Writing, review & editing. C.R.: Clinical interpretation, Writing, review & editing. S.G.: Clinical interpretation, Writing, review & editing. L.-P.N.: Conceptualization, Supervision, Funding acquisition, Writing, review & editing.

## Notes

### Author Declarations

Ethics committee/IRB of Canadian SHIELD Ethics Review Board gave ethical approval for this work (REB Tracking Number: 2023-11-003; OHRP Registration IORG0003491; FDA Registration IRB00004157; initial approval granted December 15, 2020; continuing review approval granted April 30, 2025, valid through April 29, 2026). All participants provided written informed consent prior to enrollment.

### Summary of Updates

This version updates the manuscript in three main respects. First, the author list has been expanded from three to five authors. Two co-authors have been added to reflect their contributions to the work: Caroline Rheaume (Universite Laval, Quebec City, Canada), who contributed clinical interpretation and critical revision of the manuscript, and Stephen Grobmyer (Oncology Institute, Fatima bint Mubarak Center, Cleveland Clinic Abu Dhabi, United Arab Emirates), who contributed clinical interpretation and critical revision. Author affiliations and CRediT contribution statements have been updated accordingly. Second, the manuscript text has been revised to incorporate the comments and tracked changes of the external clinical reviewers. These edits clarify the methods and improve the precision of several statements. In particular, the wording describing panel selection leakage has been harmonized to a more conservative formulation across the abstract, contributions, methods and limitations: claims that the design eliminates panel selection leakage by construction have been replaced by statements that the design minimizes panel selection leakage in the cross validation estimates. Reviewer queries that appeared inline in the draft have been removed, a small number of typographical errors have been corrected, and minor editorial improvements have been made for clarity. Third, the competing interests statement has been updated to reflect the expanded author list: it now specifies that Nicolas Anctil, Pierrick Hauguel and Louis-Philippe Noel are employees and shareholders of BioTwin Inc., while Caroline Rheaume and Stephen Grobmyer are external academic and clinical collaborators who declare no financial or ownership interest in BioTwin Inc. The study cohort, metabolomic data, analytical methods, classifier models and quantitative results are unchanged from the previous version. No new data, analyses or figures have been added, and the conclusions are unchanged.

